# Early Detection and Prediction of Emerging and Re-emerging Infectious Diseases using Data-driven Modelling: A focus on the 2022-2024 Global Monkeypox Viral Outbreak

**DOI:** 10.64898/2026.01.26.26344832

**Authors:** Thomas Wanyama James, Apellas Abaho, Simon Bbumba, Augustine Hakiza, Fortunate Amanya

**Affiliations:** Department of Plant Sciences, Microbiology & Biotechnology, College of Natural Sciences, Makerere University, P.O. Box 7062, Kampala, Uganda; Faculty of Biological and Veterinary Sciences, Nicolaus Copernicus University in Torun, Poland; Department of Chemistry, College of Natural Sciences, Makerere University, P.O. Box 7062, Kampala, Uganda; Research Department, Uganda Wildlife Research and Training Institute, P.O. BOX 173, Kasese, Uganda; Department of Records and Archives Management, College of Computing and Information Sciences, Makerere University, P.O. Box 7062, Kampala, Uganda

**Keywords:** deep neural network, data-driven modeling, gradient boosting, global monkeypox viral outbreak, polynomial regression, emerging and re-emerging infectious diseases

## Abstract

Monkeypox viral disease has been and continues to be a global public health concern. Currently, there are existing, though minimal measures to manage mpox and any future outbreaks. Relying on data-driven modeling for early detection of mpox and prediction of possible cases and deaths in the presence of an outbreak is thus imperative. The present study forecasted global mpox virus cases and deaths in Asia, Africa, Australia, Europe, North America, Oceania, and South America. Three forecasting models (deep neural network, gradient boosting, and polynomial regression) were trained on data from the seven geographical regions. The performance of the three models was assessed using coefficient of determination, mean squared error, root mean squared error, and mean absolute scaled error across each region. Prediction using the deep neural network revealed a potential of higher mpox deaths in Africa and higher mpox cases in South America. Prediction using gradient boosting showed a potential of mpox deaths in Africa and higher mpox cases in Asia and North America. Prediction using polynomial regression revealed a potential of higher mpox deaths in Africa and Asia while rapid rises in mpox cases from 2025 to 2028 were anticipated in all regions except Asia in case of a monkeypox outbreak. For the three models, the tree-based ML model (gradient boosting) outperformed the statistical model and deep learning model by R^2^ and MSE in predicting mpox case counts across all the seven geographical regions. This study showcases the worth in using data-driven modelling to predict emerging and re-emerging infectious diseases such as mpox.

## 1. Introduction

Monkeypox is one of the many global zoonotic viruses [1]. The monkeypox disease presents with symptoms like muscle pain, headache, fever, and a rash that starts as small bumps and evolves into raised bumps filled with fluid [2]. In extreme cases, the monkeypox infection may lead to dangerous complications such as pneumonia and sepsis, which can potentially become fatal [3]. Other than the monkeypox virus, other most well-known viruses responsible for epidemics include Ebola, severe acute respiratory syndrome corona virus-2 (SARS-CoV-2), Human Immunodeficiency virus-1 (HIV-1), Influenza, small pox, and severe acute respiratory syndrome (SARS) [3].

The monkeypox virus, which is the etiological agent of the monkeypox (mpox) disease [4], is also a double-stranded DNA virus belonging to Orthopoxvirus genus within the Poxviridae family and Chordopoxvirinae as the subfamily [5]. Other members of the Orthopoxvirus genus include Ectromelia virus, Taterapox virus, Camelpox virus, Vaccinia virus, Variola virus, and Cowpox virus [2]. The major hosts of poxviruses are rabbits, rodents, and non-human primates, which can occasionally be transmitted to humans, facilitating the occurrence of human-to-human transmission [6]. The virus can spread to humans through close contact with bodily fluids from a carrier, such as skin lesions, mucus, or saliva [3].

The occurrence of mpox (since 2017) outside endemic regions has increased, and the epidemiological profile of the disease within endemic regions has changed [7]. This may have led to the mpox emergence and re-emergence in endemic countries in 2022 [2]. Mpox has emerged and re-emerged for over five decades and yet not much is known about its virological profile and the characteristics of the disease it causes [4]. Its public health importance thus cannot be underestimated, as the potential for person-to-person transmission is an issue not only among household residents but also among caregivers of sick individuals [8].

It is important to highlight that persons living in forested regions, taking care of infected animals and in close proximity to infected individuals within the infectious period (21 days) are at increased risk of getting infected with mpox [9]. The reservoir host of mpox remains unknown, the viral, host, and environmental factors that modulate the virus maintenance in the wild, animal-to-animal transmission, zoonotic transmission and reverse spillover are still a mystery [4]. Diagnosis has been considered as being paramount to the detection and keeping monkeypox under control [8]. Other studies have also reported contact tracing as being one of the crucial ways of controlling the spread of mpox [10].

Although there is no specific vaccine for mpox [11], the smallpox vaccines have been reported to offer 85% cross-immunity against mpox [5]. However, these smallpox vaccines are not always effective [3]. It has also been reported that practicing proper hygiene, such as washing one’s hands regularly, avoiding close contact with those who are ill, and abstaining from contact with affected animals, can help to avoid contracting monkeypox [12]. Despite there being existing measures to manage mpox and any future outbreaks, these measures may not be fully effective for the early detection of the disease, as they may present some back-and-forth challenges. Therefore, considering the utilization of data-driven modeling in early detection of mpox and prediction of possible cases and deaths in the presence of an outbreak becomes indispensable.

Data-driven modeling involves the use of machine learning (ML) and deep learning (DL) algorithms, which have been praised because of their immense importance in the context of early detection of diseases [13]. For example, algorithms such as decision trees, random forests, and, support vector machines have been used to predict internationally important diseases such as COVID-19 [14]. Other machine learning algorithms such as supervised ML algorithms have been used in the prediction of infectious diseases [15]. Understanding the contribution of data-driven modeling in the early detection of mpox can help prioritize and garner measures for combatting or managing any future outbreaks of mpox. The present study thus found it plausible to predict the next probable mpox virus cases and deaths using three infectious disease forecasting methods (a statistical model, a deep learning model, and a ML based ensemble model) in seven different geographical regions which are; Asia, Africa, Australia, Europe, North America, Oceania, and South America. The two algorithms considered were; gradient boosting (GB), and deep neural network (DNN) while the statistical model was polynomial regression (PR). The study also evaluated the performance of the statistical model and two ML algorithms.

## 2. Materials and Methods

### 2.1 Dataset and its features

The dataset used in the present study was obtained from an online repository which is accessible at https://www.kaggle.com/datasets/rajatkumar30/monkeypox. The features of the original dataset are shown in table 1.

**Table 1.** Features in the original dataset.

| S/n | Features in the original dataset | Description |
| --- | --- | --- |
| 1 | Location | The geographical area or country where the data were recorded |
| 2 | date of record | The date of the record |
| 3 | iso_code | The ISO country code corresponding to the location |
| 4 | total_cases | The cumulative number of reported monkeypox cases up to the given date |
| 5 | total_deaths | The cumulative number of reported deaths due to monkeypox up to the given date |
| 6 | new_cases | The number of new monkeypox cases reported on the specific date. |
| 7 | new_deaths | The number of new deaths reported on the specific date. |
| 8 | new_cases_smoothed | The smoothed number of new cases, adjusted to account for reporting delays and trends. |
| 9 | new_deaths_smoothed | The smoothed number of new deaths, adjusted for reporting delays and trends. |
| 10 | new_cases_per_million | The number of new cases per million people in the location. |
| 11 | total_cases_per_million | The cumulative number of cases per million people in the location. |
| 12 | new_cases_smoothed_per_million | The smoothed number of new cases per million people, adjusted for trends and reporting delays. |
| 13 | new_deaths_per_million | The number of new deaths per million people in the location. |
| 14 | total_deaths_per_million | The cumulative number of deaths per million people in the location. |
| 15 | new_deaths_smoothed_per_million. | The smoothed number of new deaths per million people, adjusted for trends and reporting delays. |

### 2.2 Data cleaning

Cleaning of the dataset was done using Matlab’s data cleaning tool. After data cleaning and sorting, only four columns out of the original fifteen were retained. These columns had data on location (taken at the continental level), date of recording mpox cases and deaths, total cases, and total deaths of mpox.

### 2.3 Training of predictive models

Data on the date of recording mpox cases and deaths were extracted before the learning algorithms were trained. Feature matrices and target vectors were then created and this was followed by splitting the data into training and testing sets. 80% of the data were incorporated into the training set, and 20% added to the testing set. The three models considered were gradient boosting, deep neural network (DNN), and polynomial regression (PR). All the three models were trained in Matlab R2024a (http://www.mathworks.com/).

### 2.4 Prediction of future Monkeypox cases and deaths

Forecasting of the next mpox cases and deaths was done for the next four years from 2024, when the 2022-2024 mpox outbreak was last recorded.

### 2.5 Evaluation of forecasting models

The three models were evaluated using Matlab R2024a to assess their performance using four performance metrics. These four performance metrics were; coefficient of determination (R^2^), mean squared error (MSE), root mean squared error (RMSE), and mean absolute scaled error (MASE).

### 2.6 Data Analysis and Visualization

Descriptive statistics were performed on the historical monkeypox cases and deaths.

All graphical representations were performed on Matlab R2024a (http://www.mathworks.com/).

## 3. Results

This section presents the historical data on mpox cases and deaths from 2022 to 2024 in seven geographical regions all over the globe. These were Asia, Africa, Australia, Europe, North America, Oceania, and South America. The section goes ahead to present the likely number of mpox cases and deaths in the next four years from when the mpox outbreak was last registered in the seven geographical regions. The performance of each model is also shown.

### 3.1 Historical Monkeypox cases and deaths

The mean infection mpox case counts of mpox from 2022 to 2024 by geographical region were high in North America (i.e. 3784 cases), nearly three times the next highest mean infection case counts in Europe (Table 2). The mean death counts of mpox from 2022 to 2024 by geographical region were high in North America (i.e. 9 cases), nearly twice the next highest mean deaths counts in South America (Table 2).

**Table 2:** The total historical case and death counts for monkeypox by geographical region for 2022 to 2024, including the mean, minimum, and maximum of the regional cases and deaths.

|  | Mpox case counts |  |  | Mpox death counts |  |  |
| --- | --- | --- | --- | --- | --- | --- |
|  | Mean | Min-Max | Total | Mean | Min-Max | Total |
| **Asia** | 227.79 | 0-4494 | 2,899,712 | 0.49 | 0-21 | 6,239 |
| **Africa** | 326.84 | 1-4394 | 2,954,323 | 3.45 | 0-36 | 31,189 |
| **Australia** | 146.10 | 2-334 | 116,882 | 0.00 | 0-0 | 0 |
| **Europe** | 1191.46 | 0-27199 | 36,442,069 | 0.30 | 0-10 | 9,210 |
| **North America** | 3783.49 | 1-39997 | 51,270,105 | 8.59 | 0-97 | 116,464 |
| **Oceania** | 92.79 | 1-389 | 175,271 | 0.0 | 0-0 | 0 |
| **South America** | 2980.29 | 1-22915 | 28,664,454 | 5.00 | 0-44 | 48,108 |

### 3.2 Future global monkeypox cases and deaths

#### 3.2.1 Forecasted global monkeypox cases and deaths using deep neural networks

The predicted global mpox cases and deaths in Africa, Asia, Australia, North America, Oceania, and South America are presented in the figures 2, 3, 4, 5, 6, 7, and 8. The predicted mpox deaths are anticipated to be high in Africa from 2025 to 2028 (Figure 2) compared to other geographical regions in case of a monkeypox outbreak. The predicted mpox cases are expected to be high in South America in 2025 up to 2028 (Figure 8) compared to other geographical regions in case of a monkeypox outbreak. It is also key to highlight that mpox cases are anticipated to be extremely low in Australia (Figure 4) compared to other geographical regions in case of a monkeypox outbreak.

**Fig 1.**
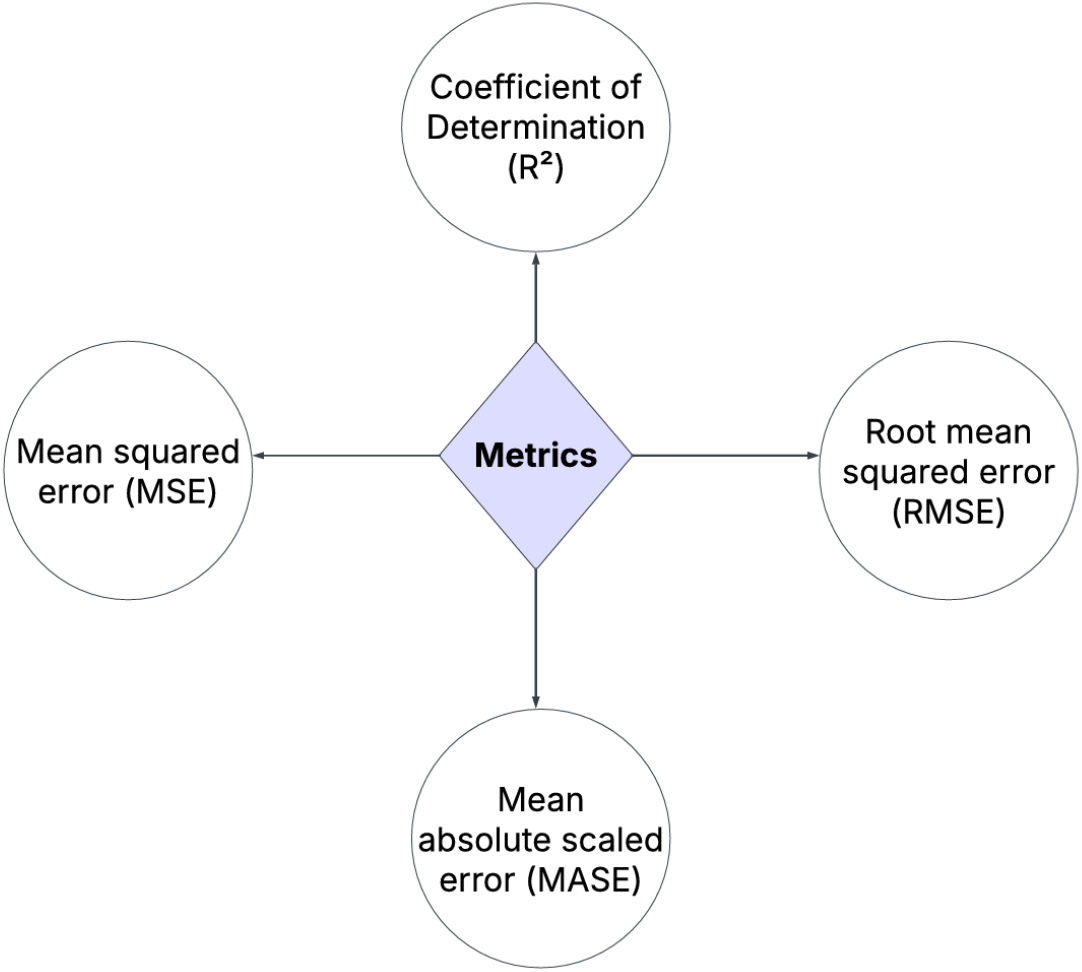
Metrics used to evaluate the performance of predictive algorithms

**Fig 2.**
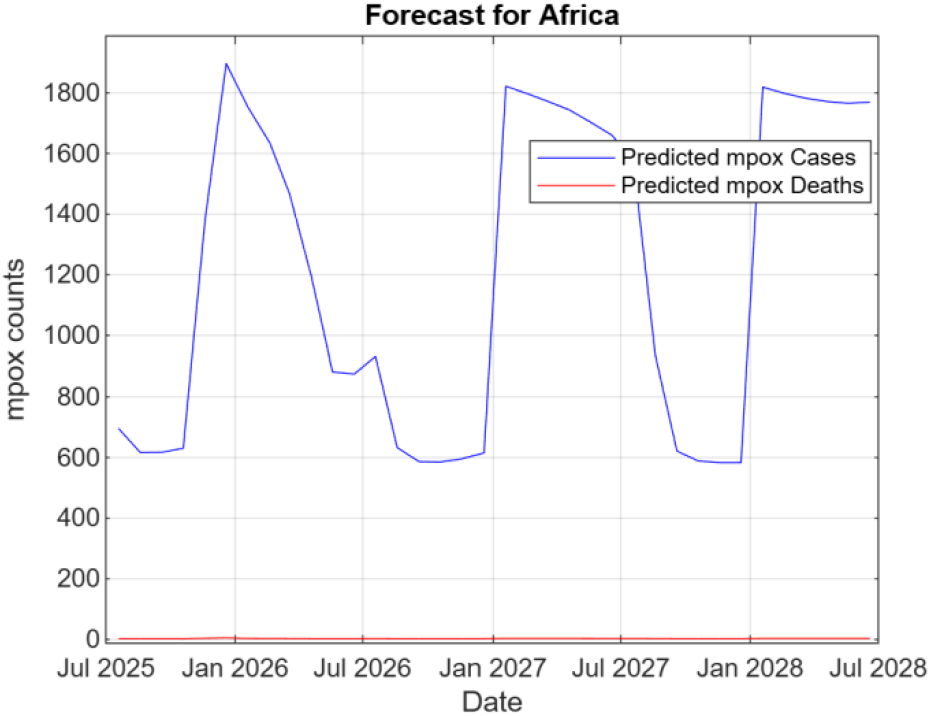
Predicted mpox cases and deaths in Africa using DNN

**Fig 3.**
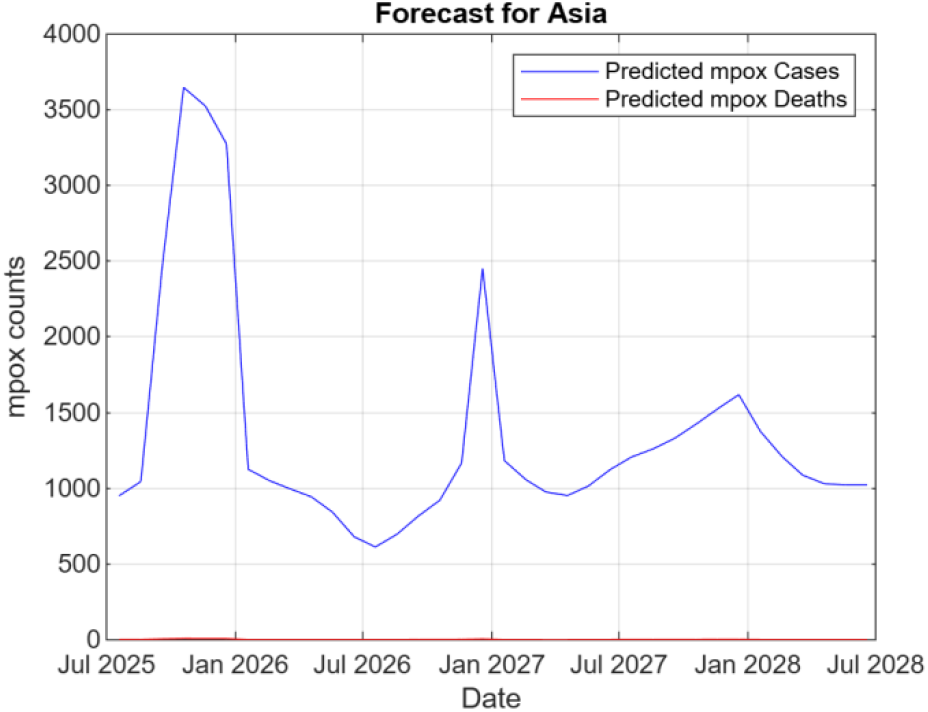
Predicted mpox cases and deaths in Asia using DNN

**Fig 4.**
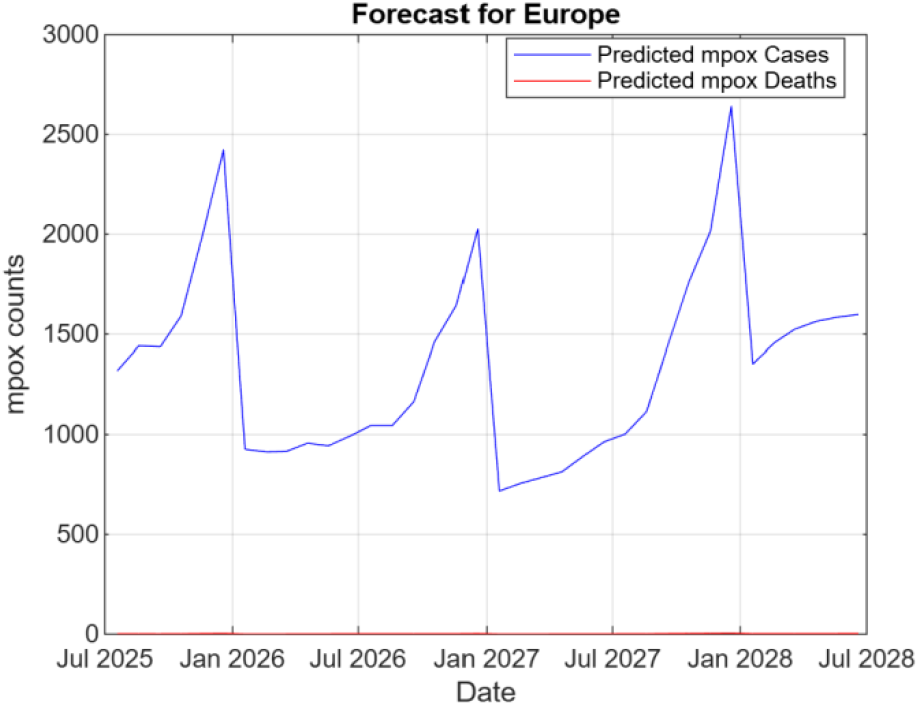
Predicted mpox cases and deaths in Europe using DNN

**Fig 5.**
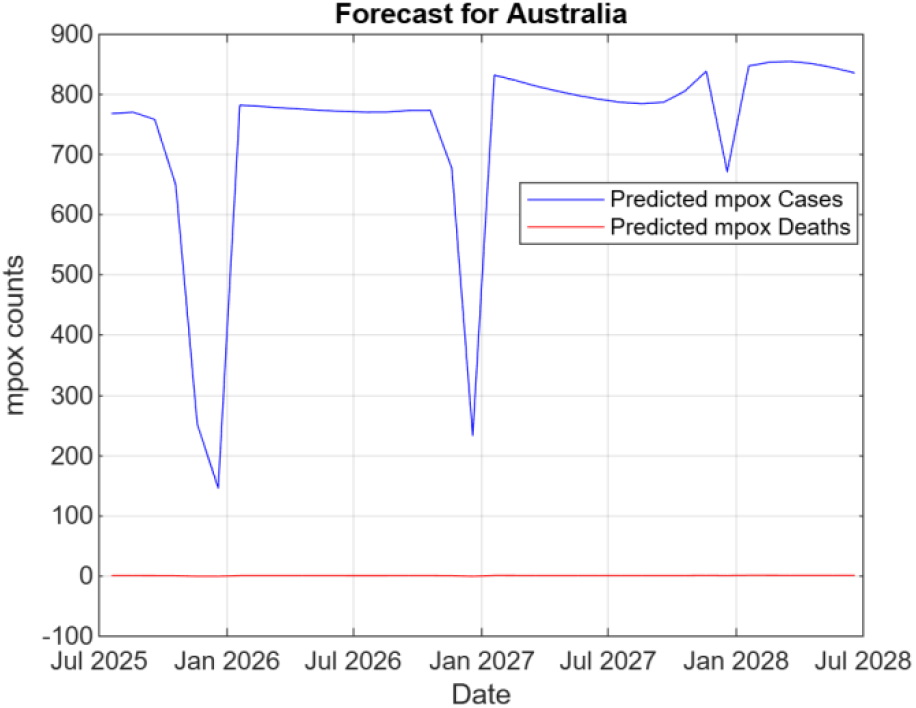
Predicted mpox cases and deaths in Australia using DNN

**Fig 6.**
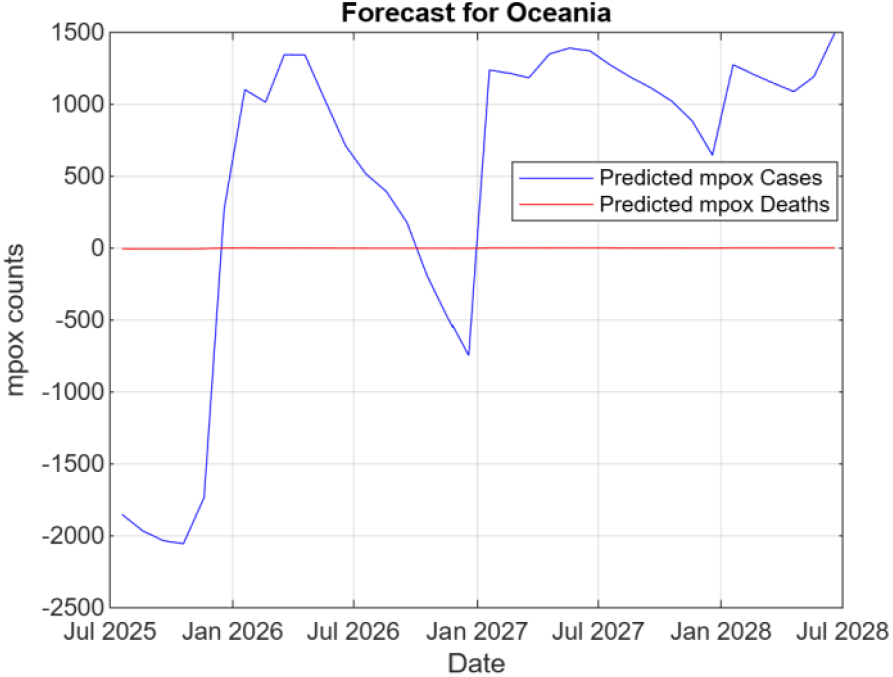
Predicted mpox cases and deaths in Oceania using DNN

**Fig 7.**
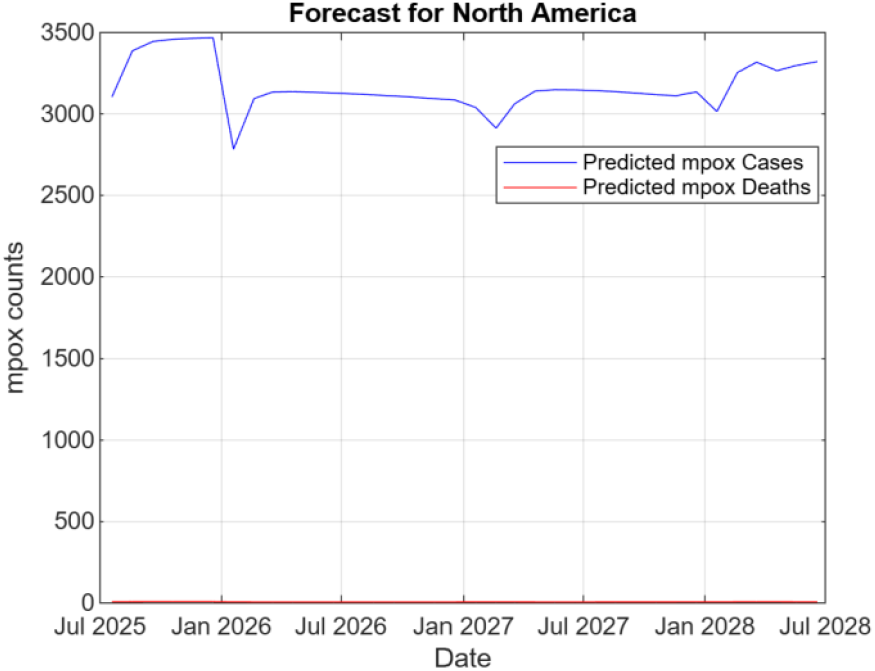
Predicted mpox cases and deaths in North America using DNN

**Fig 8.**
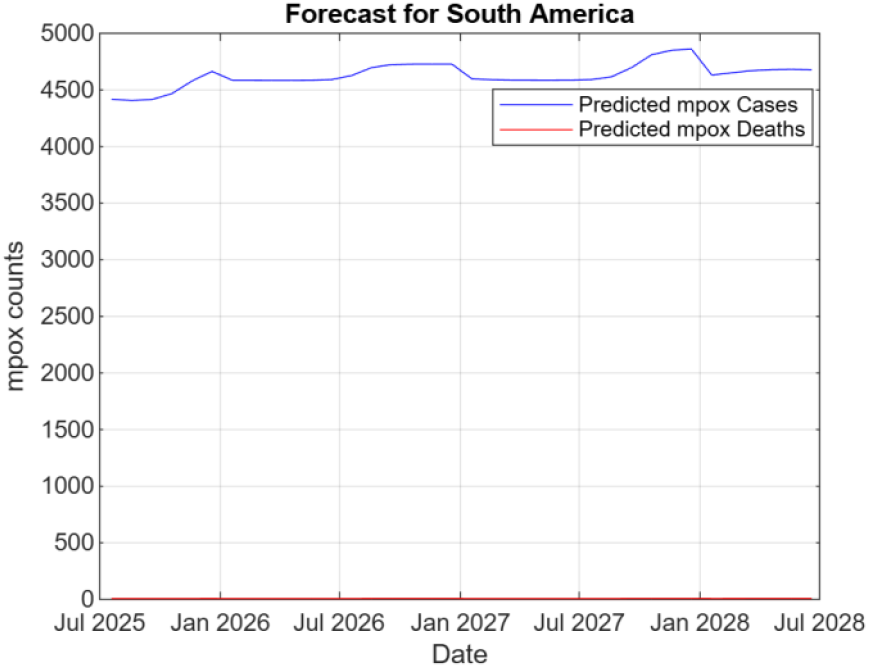
Predicted mpox cases and deaths in South America using DNN

#### 3.2.2. Forecasted global monkeypox cases and deaths using gradient boosting

The predicted global mpox cases and deaths in Africa, Asia, Australia, North America, Oceania, and South America are presented in the figures 9, 10, 11, 12, 13, 14, and 15. The global monkeypox death counts are anticipated to be high in Africa (figure 9) compared to other geographical regions in case of a monkeypox outbreak. Monkeypox cases are anticipated to be high in Asia (figure 10) and North America (figure 13) compared to other geographical regions in case of a monkeypox outbreak. Monkeypox cases are expected to be extremely low in Australia (figure 11) and Oceania (figure 14) compared to other geographical regions in case of a monkeypox outbreak.

**Fig 9.**
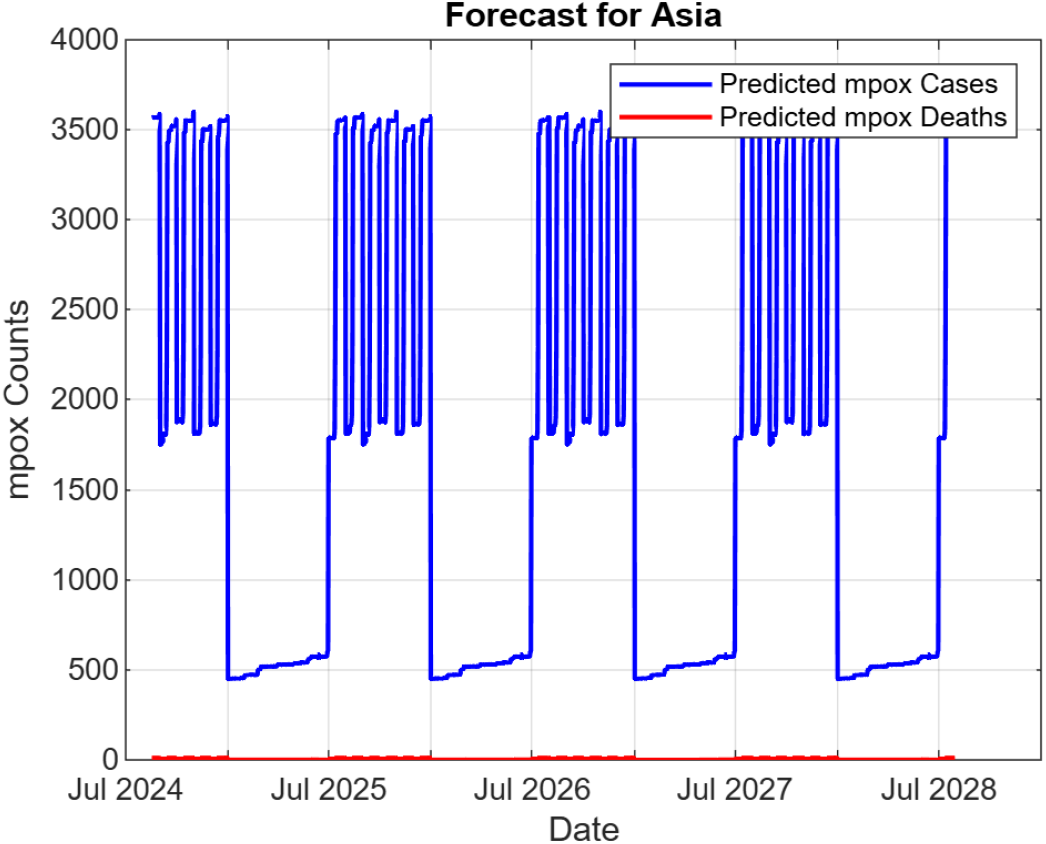
Predicted mpox cases and deaths in Asia using GB

**Fig 10.**
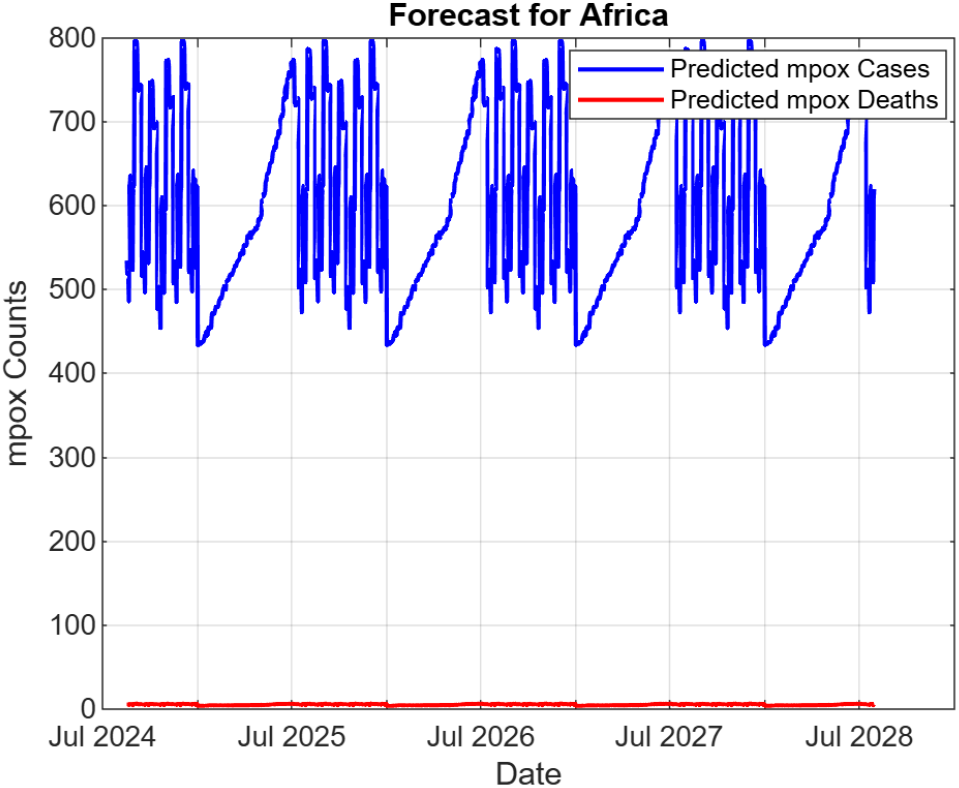
Predicted mpox cases and deaths in Africa using GB

**Fig 11.**
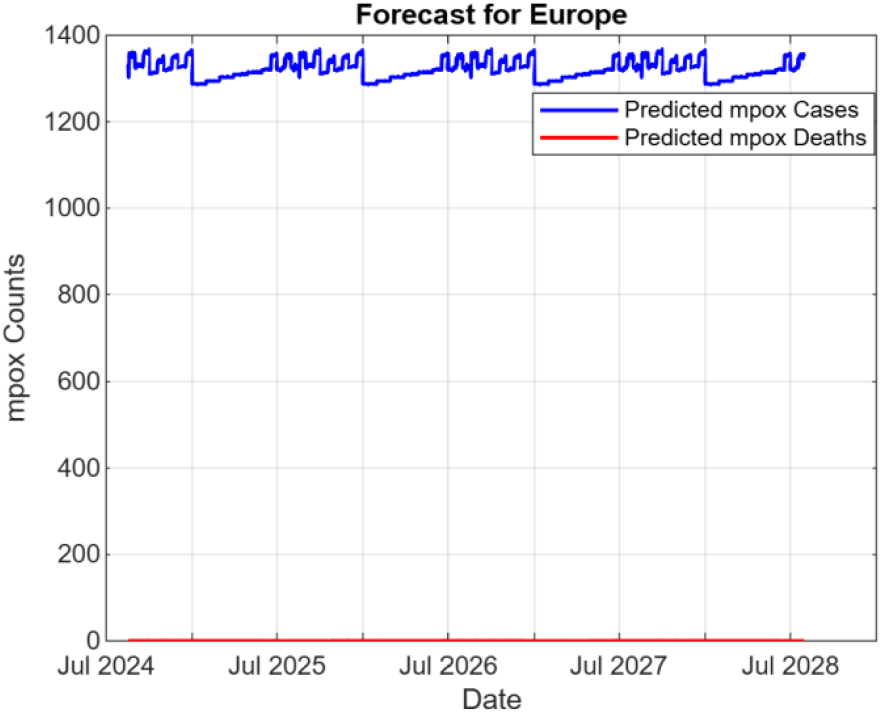
Predicted mpox cases and deaths in Europe using GB

**Fig 12.**
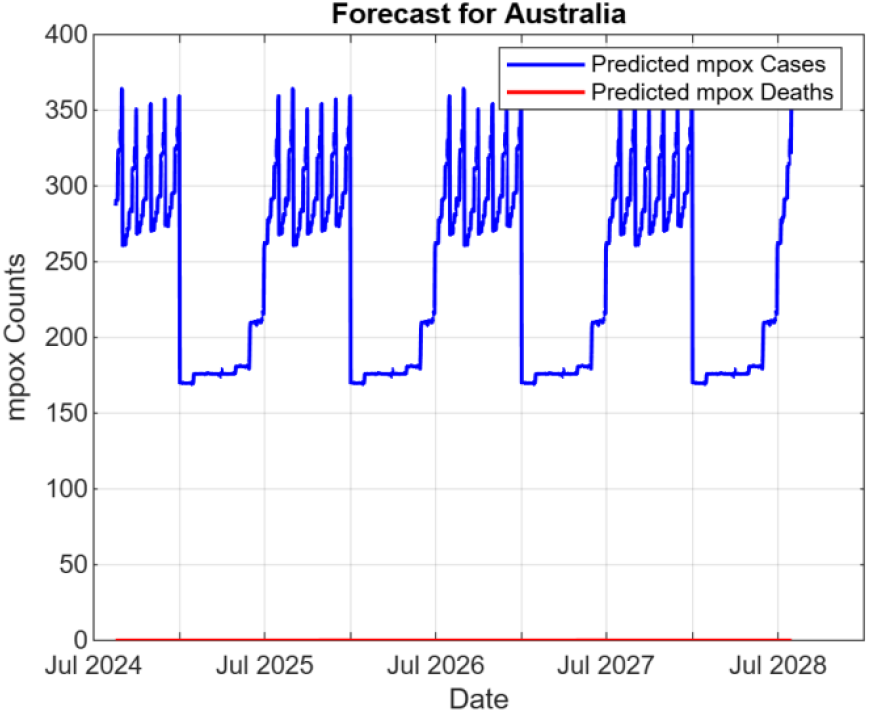
Predicted mpox cases and deaths in Australia using GB

**Fig 13.**
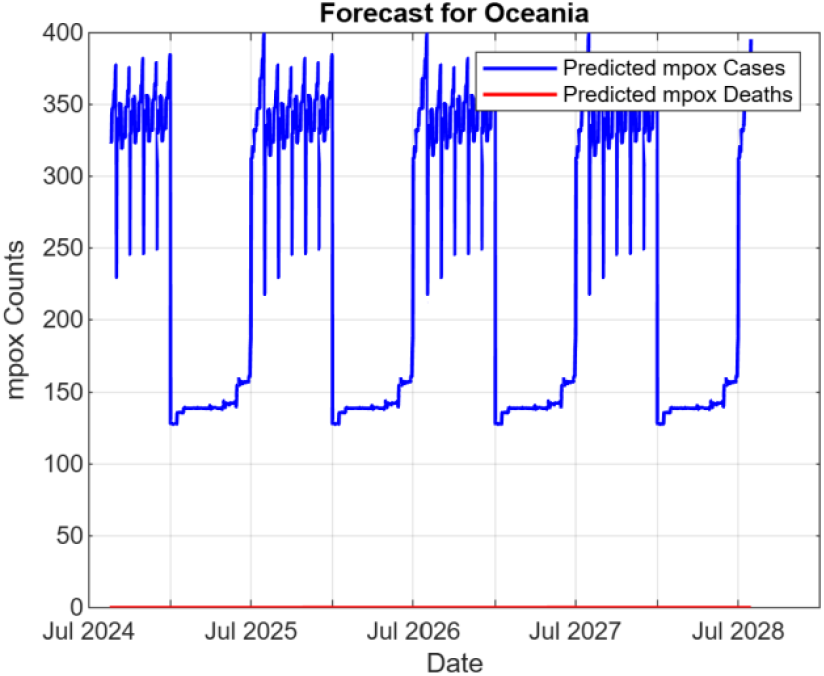
Predicted mpox cases and deaths in Oceania using GB

**Fig 14.**
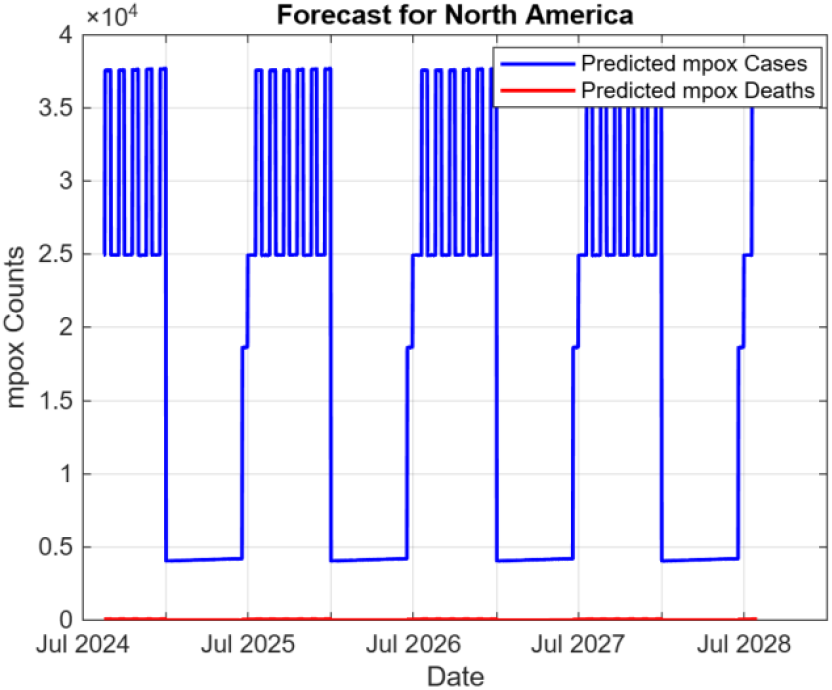
Predicted mpox cases and deaths in North America using GB

**Fig 15.**
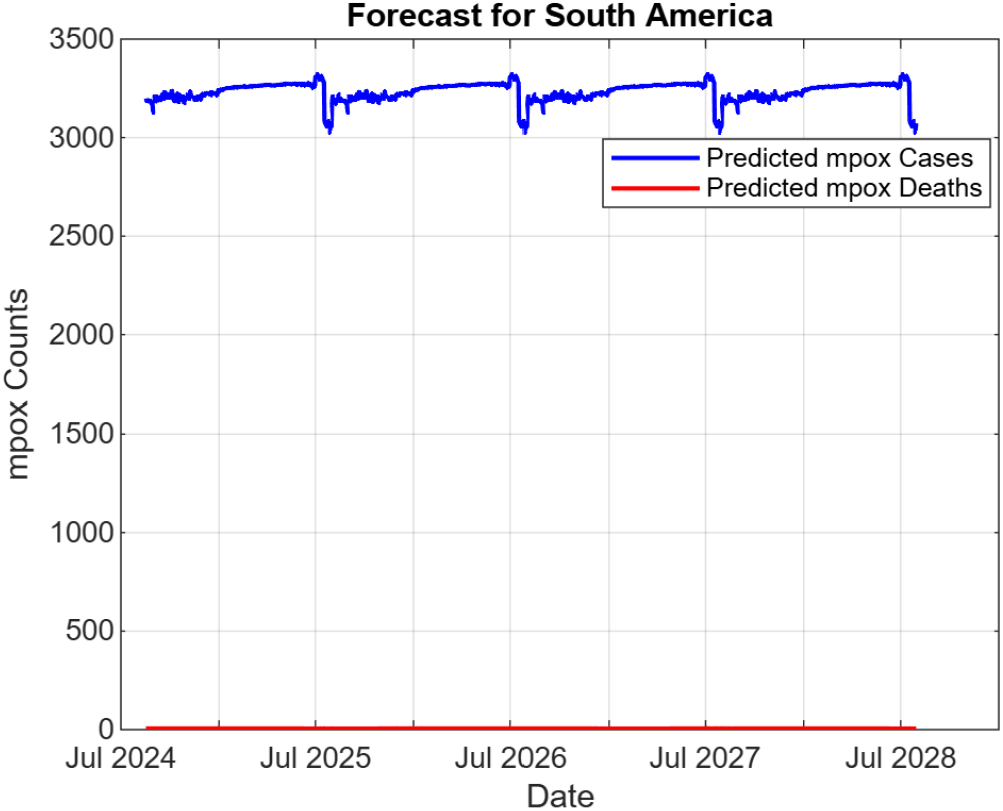
Predicted mpox cases and deaths in South America using GB

#### 3.2.3 Forecasted monkeypox cases and deaths using polynomial regression

The predicted global mpox cases and deaths in Africa, Asia, Australia, North America, Oceania, and South America are presented in the figures 16, 17, 18, 19, 20, 21, and 22. In case of a monkeypox outbreak, the global mpox death counts are expected to increase from 2024 to 2028 in Africa (figure 16) and Asia (figure 17). However, the global mpox death counts are likely to remain constant from 2024 to 2028 in Australia (figure 18), Europe (figure 19), North America (figure 20), Oceania (figure 21), and South America (figure 22) in the presence of an outbreak. The global mpox case counts are anticipated to rapidly rise from 2024 to 2028 in Asia (figure 17). Sharp rises are also expected to occur from 2025 to 2028 in Africa (figure 16), Australia (figure 18), Europe (figure 19), North America (figure 20), Oceania (figure 21), and South America (figure 22) in case of a monkeypox outbreak.

**Fig 16.**
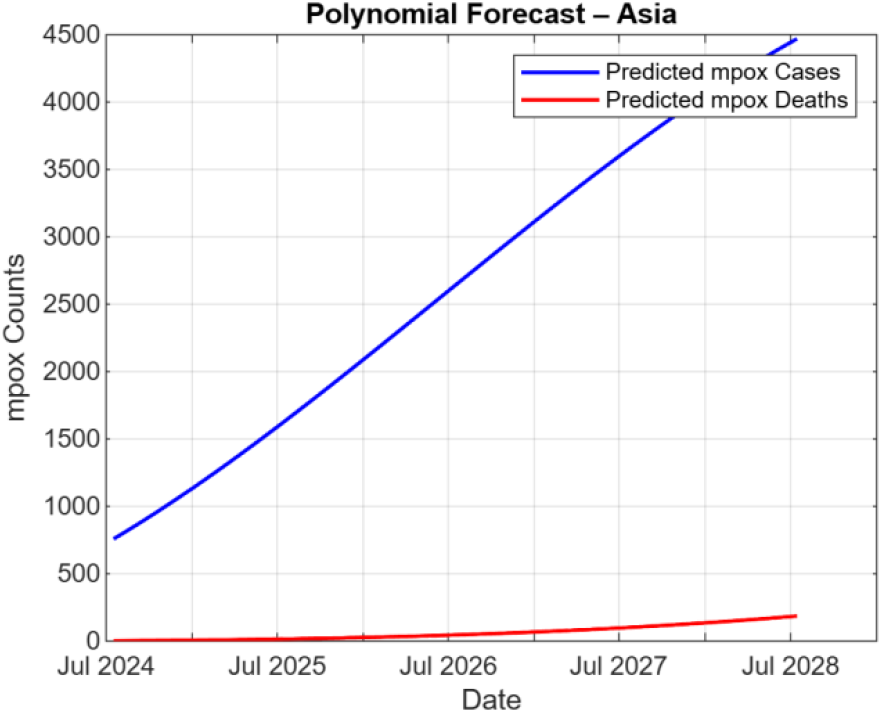
Predicted mpox cases and deaths in Asia using PR

**Fig 17.**
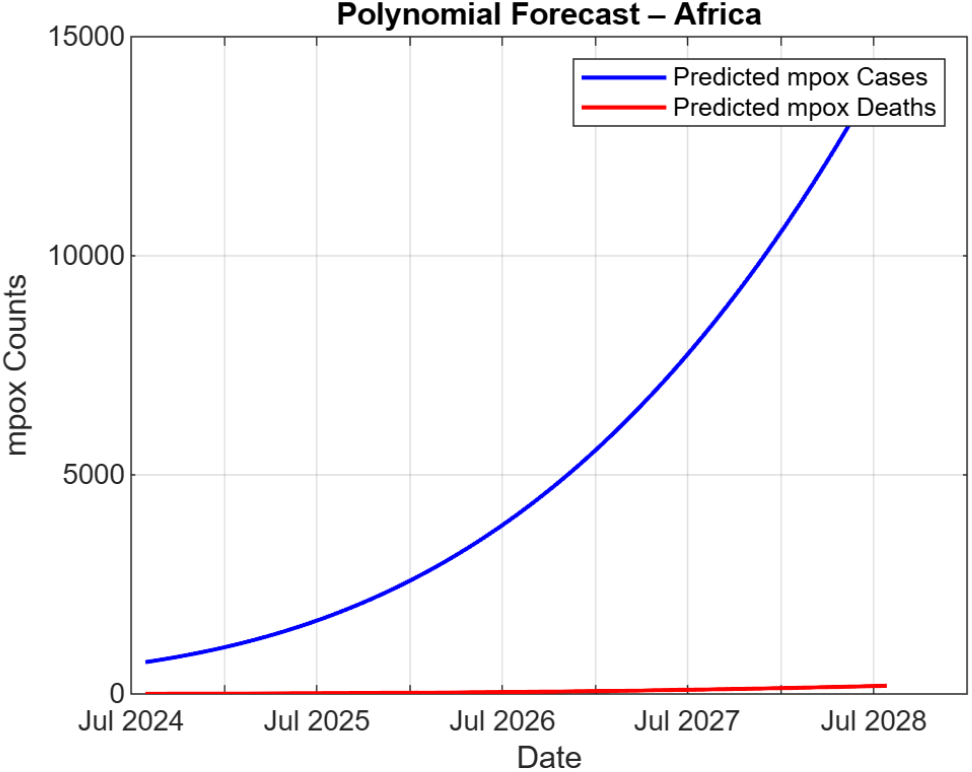
Predicted mpox cases and deaths in Africa using PR

**Fig 18.**
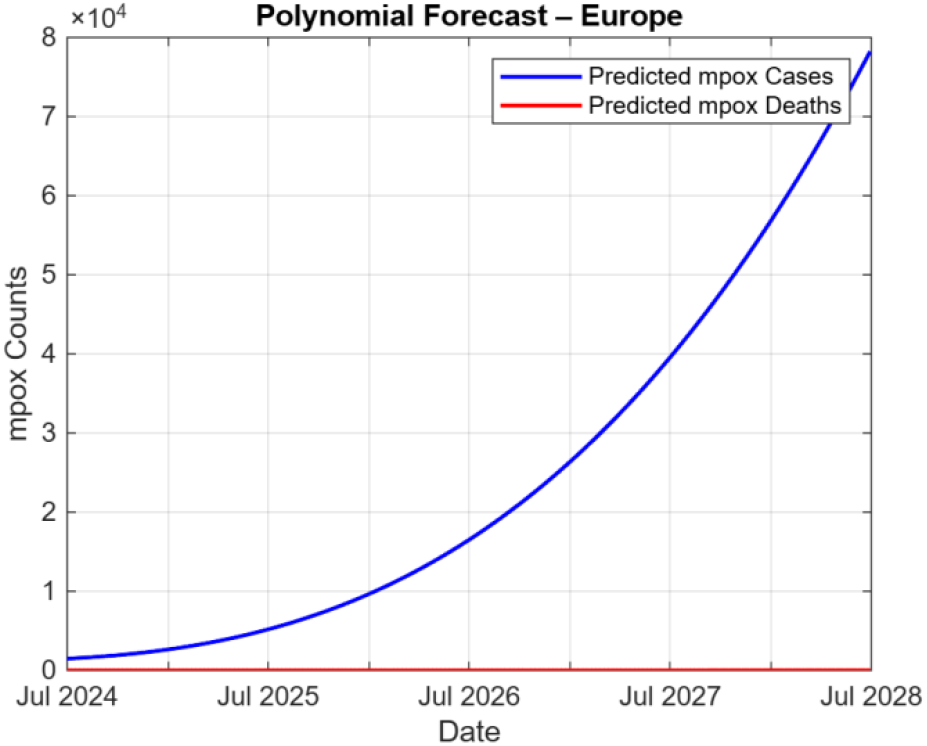
Predicted mpox cases and deaths in Europe using PR

**Fig 19.**
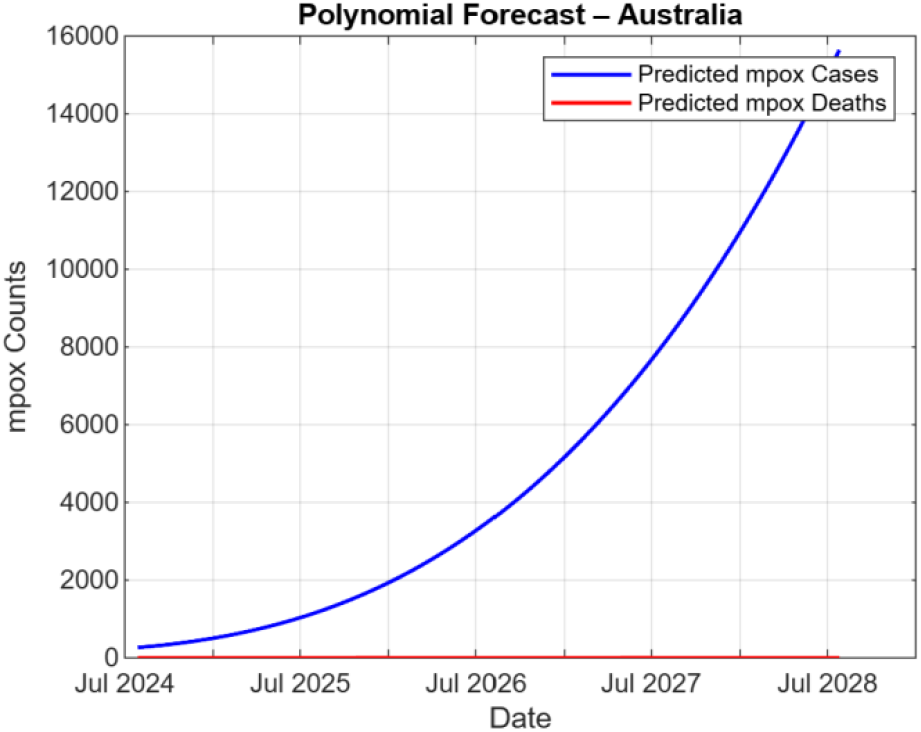
Predicted mpox cases and deaths in Australia using PR

**Fig 20.**
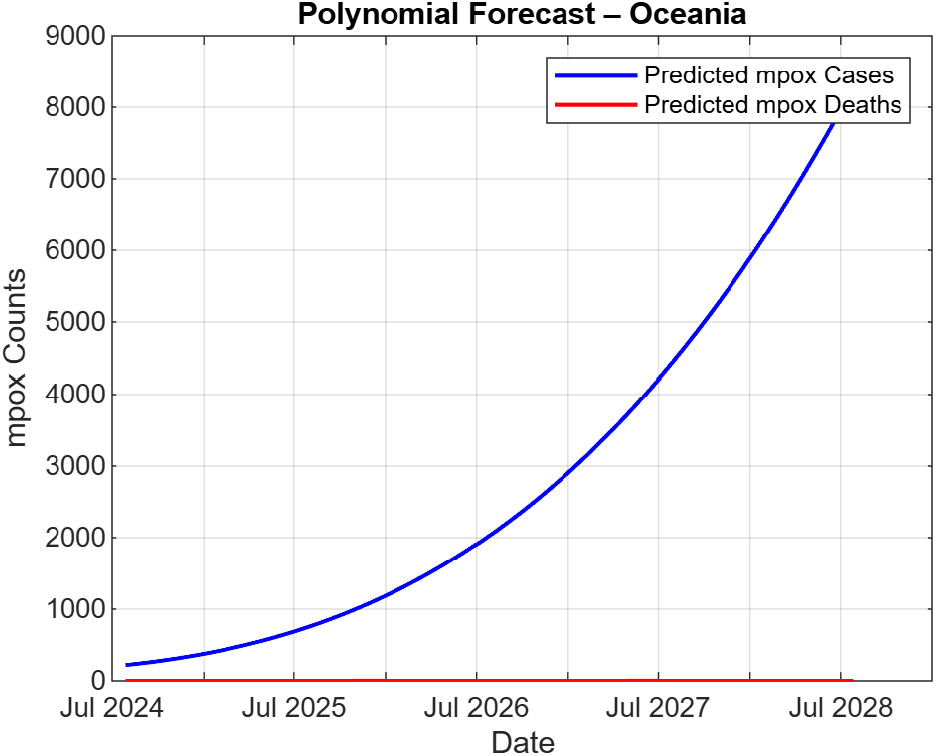
Predicted mpox cases and deaths in Oceania using PR

**Fig 21.**
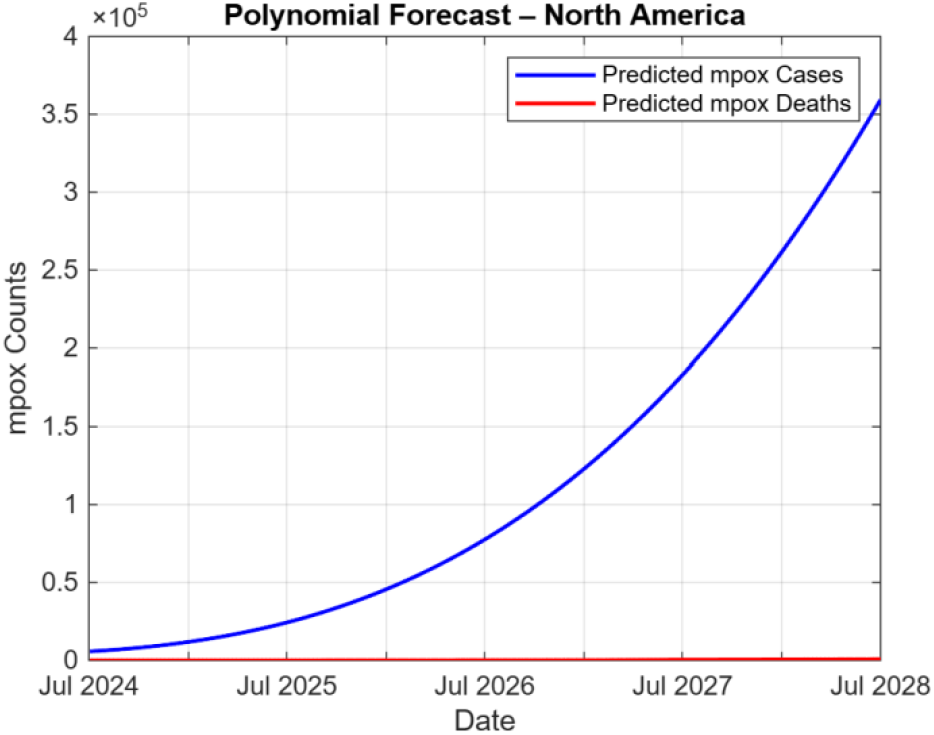
Predicted mpox cases and deaths in North America using PR

**Fig 22.**
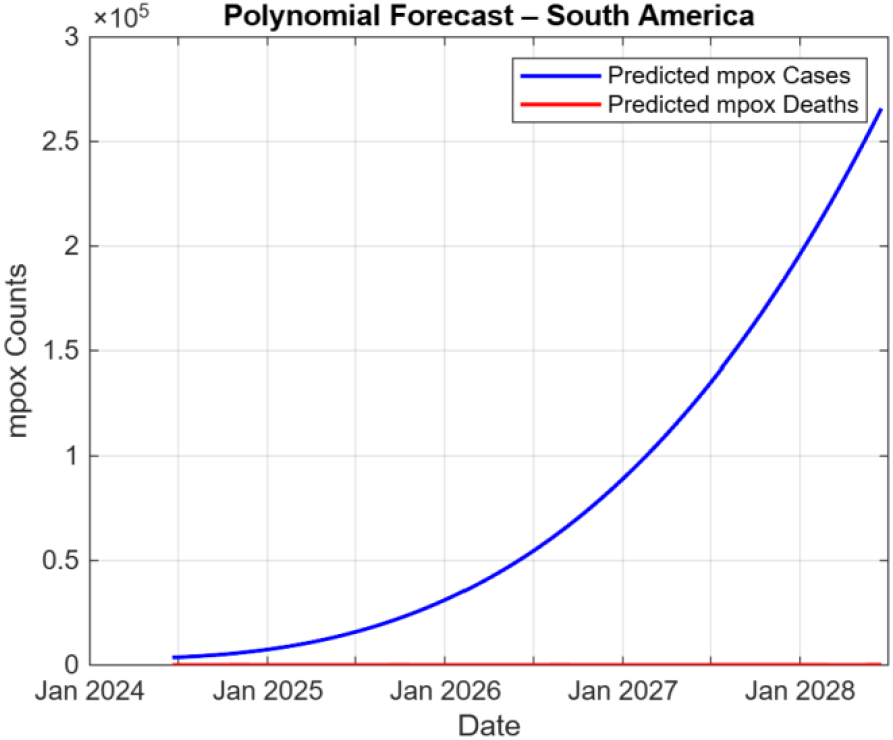
Predicted mpox cases and deaths in South America using PR

### 3.2 Performance of forecasting models by geographical region

The performance of each model by geographical region is presented in Tables 3, 4, and 5.

**Table 3.** Performance metrics of DNN by geographical region.

| Geographical region | Mpox case counts |  |  |  | Mpox death counts |  |  |  |
| --- | --- | --- | --- | --- | --- | --- | --- | --- |
|  | MSE | RMSE | R<sup>2</sup> | MASE | MSE | RMSE | R<sup>2</sup> | MASE |
| **Africa** | 29635283.63 | 3684.67775 | -0.9234833 | 60.3556333 | 5880.85613 | 76.6607667 | -1489.9276 | 1.17462E+17 |
| **Asia** | 5444082 | 737.6192 | -0.0787 | 34.7065 | 6048.3013 | 77.7708 | -1395.1282 | 2634.0505 |
| **Australia** | 15734.2861 | 125.4364 | -4.6521 | 54.5512 | 6122.5591 | 78.2468 | -Inf | 352393648500899840.0000 |
| **Europe** | 17852298.0000 | 4225.1978 | -0.0854 | 70.8072 | 6074.4692 | 77.9389 | -4504.022 | 11860.9795 |
| **North America** | 117738280.0000 | 10850.7275 | -0.1339 | 81.0803 | 5380.7476 | 73.3536 | -9.9896 | 589.8707 |
| **Oceania** | 11431.4775 | 106.9181 | -0.3534 | 65.9481 | 6122.1753 | 78.2443 | -Inf | 352381347714564096.0000 |
| **South America** | 36749876.0000 | 6062.1675 | -0.2374 | 55.0405 | 5536.8843 | 74.4102 | -50.5707 | 787.4851 |

**Table 4.**
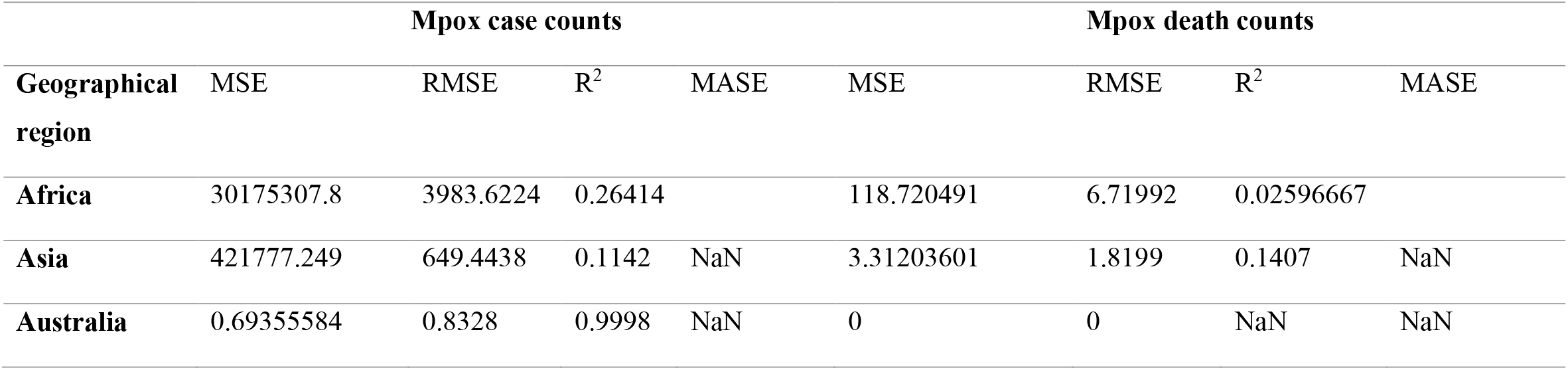
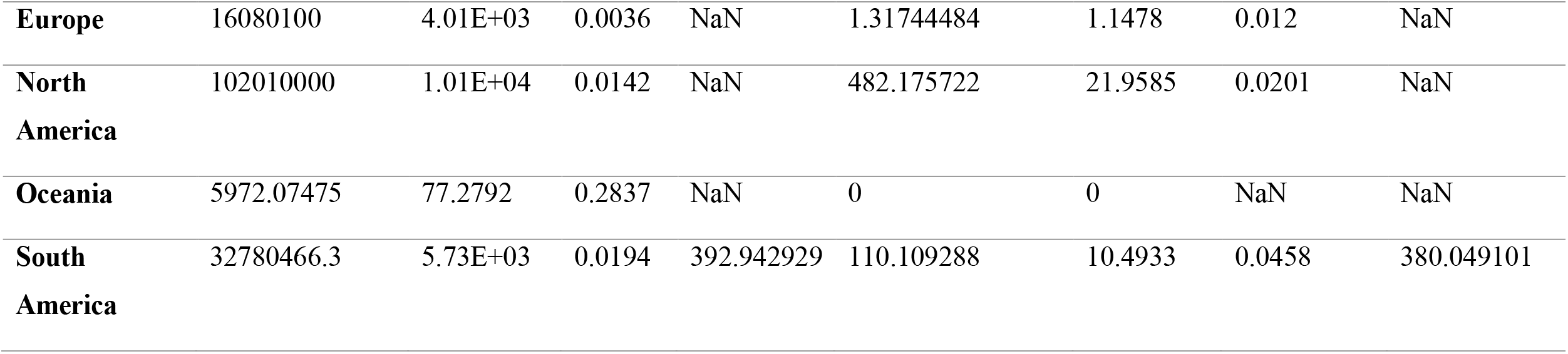
Performance metrics of gradient boosting by geographical region.

**Table 5.** Performance metrics of polynomial regression by geographical region.

| **Geographical region** | **Mpox case counts** |  |  |  | **Mpox death counts** |  |  |  |
| --- | --- | --- | --- | --- | --- | --- | --- | --- |
|  | MSE | RMSE | R<sup>2</sup> | MASE | MSE | RMSE | R<sup>2</sup> | MASE |
| **Africa** | 443798.1183 | 666.1817457 | 0.070610973 | 0.712760965 | 35.8871412 | 5.990587717 | 0.046304529 | 0.633856981 |
| **Asia** | 431874.3201 | 657.1714541 | 0.092948552 | 0.933404125 | 3.40253851 | 1.844597113 | 0.117209987 | 1.008789056 |
| **Australia** | 236.3974353 | 15.37522147 | 0.916109156 | 31.03213263 | 0 | 0 | NaN | NaN |
| **Europe** | 16078693.24 | 4009.824589 | 0.002972155 | 0.782036074 | 1.318178435 | 1.148119521 | 0.0114963 | 0.894279444 |
| **North America** | 103134576.4 | 10155.51951 | 0.005847214 | 0.869658458 | 485.743468 | 22.03958865 | 0.012830058 | 1.018746521 |
| **Oceania** | 6392.393384 | 79.95244451 | 0.233301074 | 0.572607049 | 0 | 0 | NaN | NaN |
| **South America** | 32815708.23 | 5728.499649 | 0.018356641 | 0.757258816 | 110.5597664 | 10.51474044 | 0.041907505 | 0.938118699 |

Across all geographical regions, the tree-based ML model (gradient boosting) outperformed the statistical model (polynomial regression) and deep learning model (deep neural network) by R^2^ and MSE in predicting mpox case counts. The tree-based ML model (gradient boosting) and statistical model (polynomial regression) both outperformed the deep learning model (deep neural network) by RMSE in predicting mpox cases and death counts. The statistical model outperformed the deep learning model (deep neural network) by MASE in predicting mpox cases and death counts. The tree-based ML model outperformed the statistical model and deep learning model by MSE in predicting mpox death counts. All the predictions by MASE were worse than the naïve predictions across all geographical regions.

Region wise, for the root mean squared error (RMSE) and mean squared error (MSE), deep neural networks had a better performance in predicting mpox case counts in Oceania compared to other geographical regions (Table 3). For the MASE, the deep learning model had a better performance in predicting mpox case counts in Asia and mpox death counts in North America compared to other geographical regions.

Using gradient boosting (Table 4), a good performance was observed in Australia in predicting mpox case counts by MSE, RMSE and R^2^ compared to other geographical regions. A good performance was observed in Asia in predicting mpox death counts by R^2^ and a good performance in Europe in predicting mpox death counts by RMSE and MSE.

Using polynomial regression (Table 5), a good performance in predicting mpox case counts was observed in Australia by MSE, RMSE, and R^2^ compared to other geographical regions. For the MASE, better performance in predicting mpox case counts was recorded in Oceania. For prediction of mpox death counts, a better performance by MASE was in Africa. A good performance of the statistical model by R^2^, MSE and RMSE was in Europe.

## 4. Discussion

Infectious diseases are a growing part of the global health burdens as millions of deaths are registered yearly due to infectious diseases [16]. The spread of these infectious diseases, whether emergent such as COVID-19, which has caused close to 7 million deaths globally or long-standing infectious diseases such as malaria, significantly impacts human well-being and social development on a universal scale [17]. Therefore, the fight against infectious disease is nonstop [18]. The progress by humans in coming up with measures to combat infectious diseases has greatly depended on scientific innovations across multiple disciplines [18].

One of these scientific innovations is machine learning, which has proven exceptionally effective in infectious disease modeling due to its capability to handle big data and also reveal robust patterns [18]. This ability has fuelled its widespread and fruitful application in key functions associated with understanding and battling the spread of infectious diseases [19]. Among the applications of machine learning are; prediction of drug-virus interactions [20], identification of propagation source of infectious diseases [21], detection of individual infections [22], and intervention planning [23].

Machine learning also allows for modeling and prediction of transmission risks of infectious diseases thus helping to guide public health decisions and therefore draw intervention measures [24]. The present study considered three forecasting models (deep neural network, gradient boosting, and polynomial regression) to predict mpox cases and deaths from seven geographical regions (Africa, Asia, Australia, Europe, North America, Oceania, and South America) since 2024. The dataset used was large as it contained mpox cases and deaths in different countries all over the globe.

The different countries were then categorized depending on their geographical locations, thus the seven geographical regions of Africa, Asia, Australia, Europe, North America, Oceania, and South America. Across the seven geographical regions, the DL model (deep neural network) performed poorly in prediction of both global mpox cases and deaths. This observation has also been reported in another study where the DL models (encoder–decoder and multi-layer perceptron model) used to forecast three infectious diseases (typhoid, campylobacteriosis, and Q-fever) all had a poor performance [25]. The same observation in the present study may be that the DL model used (deep neural network) suffered from over-fitting, likely due to causes such as learning the noise of the training data [26]. Despite the poor performance of the DL model used in the current study, DL models have been and continue to be praised for outperforming ML models (such as gradient boosting) in other tasks [27].

Mpox, an unusual zoonotic disease caused by an Orthopoxvirus [28], was first discovered in laboratory monkeys in 1958 and in humans in the Democratic Republic of the Congo in 1970 [29]. It was originally confined to West and Central Africa, but has now spread to other geographical regions such as Asia and Europe, thus provoking the World Health Organization to declare the disease a global public health concern in May 2022 and August 2024 [16]. The 2022 and 2024 mpox outbreaks were therefore the backbone of the present study, which focused on the early detection and prediction of mpox, an emerging and re-emerging infectious disease, using data-driven modeling.

Data-driven modeling was opted for since it has been proven effective in spotting diverse diseases [30]. The present study has several immense strengths in the field of infectious disease modeling and prediction. This is because infectious diseases are drivers of significant increases in incidence and mortality, which fuel substantial losses to society and livelihood, and bring heavy burdens to public health systems [18]. One of the strengths is that the study engineered a single dataset having two explanatory variables covering seven regions with six being continents (Africa, Asia, Europe, Australia, North America, and South America) and one geographical regions (Oceania). Each of the six continents comprised of over ten countries while Oceania had over five countries.

Three forecasting models; one being a DL model (deep neural network), the other a tree-based ML model (gradient boosting), and the last being a statistical model (polynomial regression), were applied using the same feature inputs for the monkeypox disease. These three models were also evaluated using four metrics, MSE, RMSE, R^2^, and MASE. This was done to provide a comprehensive understanding of the performance of each forecasting model.

## 5. Conclusion

This study’s aim was to forecast mpox cases and deaths in seven geographical locations. Though not the gist of the study, the mean mpox cases and deaths were computed. The mean mpox case and death counts from 2022 to 2024 were high in North America compared to other regions. Mpox cases and deaths were anticipated to increase from 2024 to 2028 basing on predictions from deep neural network, gradient boosting, and polynomial regression. After prediction from deep neural networks and gradient boosting, the mpox cases were anticipated to be extremely low in Australia compared to other geographical regions in case of a monkeypox outbreak.

The global mpox death counts are expected to increase from 2024 to 2028 in Africa and Asia in case of a monkeypox outbreak. For the three forecasting models used, the tree-based ML model outperformed the statistical model and deep learning model by R^2^ and MSE in predicting mpox case counts across all the seven geographical regions. Put together, the study showcases the application of data-driven modelling in forecasting emerging and re-emerging infectious diseases. This research puts forth that data-driven modelling is a sustainable tool that can be merged with other measures to combat infectious diseases. Therefore, utilization of predictive models can help to mitigate future monkeypox outbreaks.

## Data Availability

All data produced in the present work are contained in the manuscript

https://www.kaggle.com/datasets/rajatkumar30/monkeypox.

## Abbreviations

ML: machine Learning
DL: deep learning
DNN: deep neural network
GB: gradient boosting
PR: polynomial regression
MSE: mean squared error
RMSE: root mean squared error
MASE: mean absolute scaled error

## Author contributions

Research concept, design, and methodology: Thomas James Wanyama Data analysis: Thomas James Wanyama

Writing draft manuscript: Thomas James Wanyama

Writing, review, and editing: Simon Bbumba, Augustine Hakiza, Apellas Abaho, and Fortunate Amanya.

## Clinical trial number

Not applicable.

## Consent to Publish declaration

Not applicable.

## Consent to Participate declaration

Not applicable.

## Ethics declaration

Not applicable.

## Availability of data and materials

All the data used in the study are publicly available with website links in the Methods section.

